# CAPYBARA: A Generalizable Framework for Predicting Serological Measurements Across Human Cohorts

**DOI:** 10.1101/2025.07.07.25331040

**Authors:** Sierra Orsinelli-Rivers, Daniel Beaglehole, Tal Einav

## Abstract

The rapid growth of biological datasets presents an opportunity to leverage past studies to inform and predict outcomes in new experiments. A central challenge is to distinguish which serological patterns are universally conserved and which are specific to individual datasets. In the context of human serology studies, where antibody-virus interactions assess the strength and breadth of the antibody response and inform vaccine design, differences in cohort demographics or experimental design can markedly impact responses, yet few methods can translate these differences into the value±uncertainty of future measurements. Here, we introduce CAPYBARA, a data-driven framework that quantifies how serological relations map across datasets. As a case study, we applied CAPYBARA to 25 influenza datasets from 1997-2023 that measured vaccine or infection responses against multiple H3N2 variants using hemagglutination inhibition (HAI). To demonstrate how a subset of measurements in each study can infer the remaining data, we withheld all HAI measurements for each variant and accurately predicted them with a 2.0-fold mean absolute error—on par with experimental assay variability. Although studies with similar designs showed the best predictive power (*e*.*g*., children data are better predicted by children than adult data), predictions across age groups, between vaccination and infection studies, and across studies conducted <5 years apart showed comparable 2−3-fold accuracy. By analyzing feature importance in this interpretable model, we identified global cross-reactivity trends, enabling future serology or vaccine studies to infer broad serological responses from a small subset of measurements.

## Introduction

As biological datasets continue to expand in size and complexity, it is becoming increasingly more challenging to integrate information from prior datasets to inform and predict the outcomes of future experiments. Patterns found in one group of individuals may not apply to another group where factors such as age, exposure history, or immune state differ.^1-6^ More subtle, and often unknowable, differences in experimental methodology or batch effects may further affect which datasets can predict one another. While many studies have identified that cohorts differ in some way (*e*.*g*., children and adults show significantly different immune responses^7-11^), we lack methods that estimate how these differences translate into future measurements. Such quantitative predictions are not only the hallmark of deeply understanding a system, but they also facilitate head-to-head comparisons across studies measuring different features.

This work tackles this problem in the context of the antibody response against the rapidly evolving influenza virus, which underpins the annual vaccine selection process.^12,13^ Specifically, we consider serum hemagglutination inhibition (HAI) against multiple influenza variants, where higher HAI titers correlate with greater protection.^14-16^ While thousands of new variants (or strains) emerge each year, only a small fraction can be functionally characterized using HAI, and the variants measured often differ between studies. Critically, we still lack methods that take a person’s HAI titers against a few variants and infer their titers for other variants, which would quantify the holes in a population’s immunity that should be closed when the vaccine is next updated.

Currently available HAI datasets have several direct clinical applications. Prior work has shown that a person’s HAI against multiple strains can infer their influenza exposure history^17,18^ or help predict their response to future vaccines.^19^ Serum-virus HAI titers have been shown to be inherently low dimensional,^20,21^ where titers against some variants can infer the titers of other strains.^22,23^ As such, a new study seeking to measure HAI against numerous variants could theoretically extract these cross-reactivity relations from existing datasets, measure a minimal number of variants, and then predict the HAI of the remaining strains. One key hurdle is that cross-reactivity relations may differ with age, influenza exposure, and other immune variables. As the number of prior studies continues to increase, it is unclear *a priori* which datasets will best predict the cross-reactivity relations in another study, nor what form those relations will take.

To that end, we introduce the <u>C</u>ross-study <u>A</u>daptive <u>P</u>redictions, <u>Y</u>ielding <u>B</u>ayesian <u>A</u>ggregation with <u>R</u>ecursive <u>A</u>nalysis (CAPYBARA), a generalizable framework that efficiently selects the most predictive features within each dataset, determines their cross-reactivity relations, estimates prediction error, and then combines predictions in a new study by heavily weighing the most confident predictions. **Figure 1** provides an overview of the CAPYBARA workflow, including the feature learning process, model training, error calibration, and Bayesian combination of predictions across datasets.

**Figure 1.**
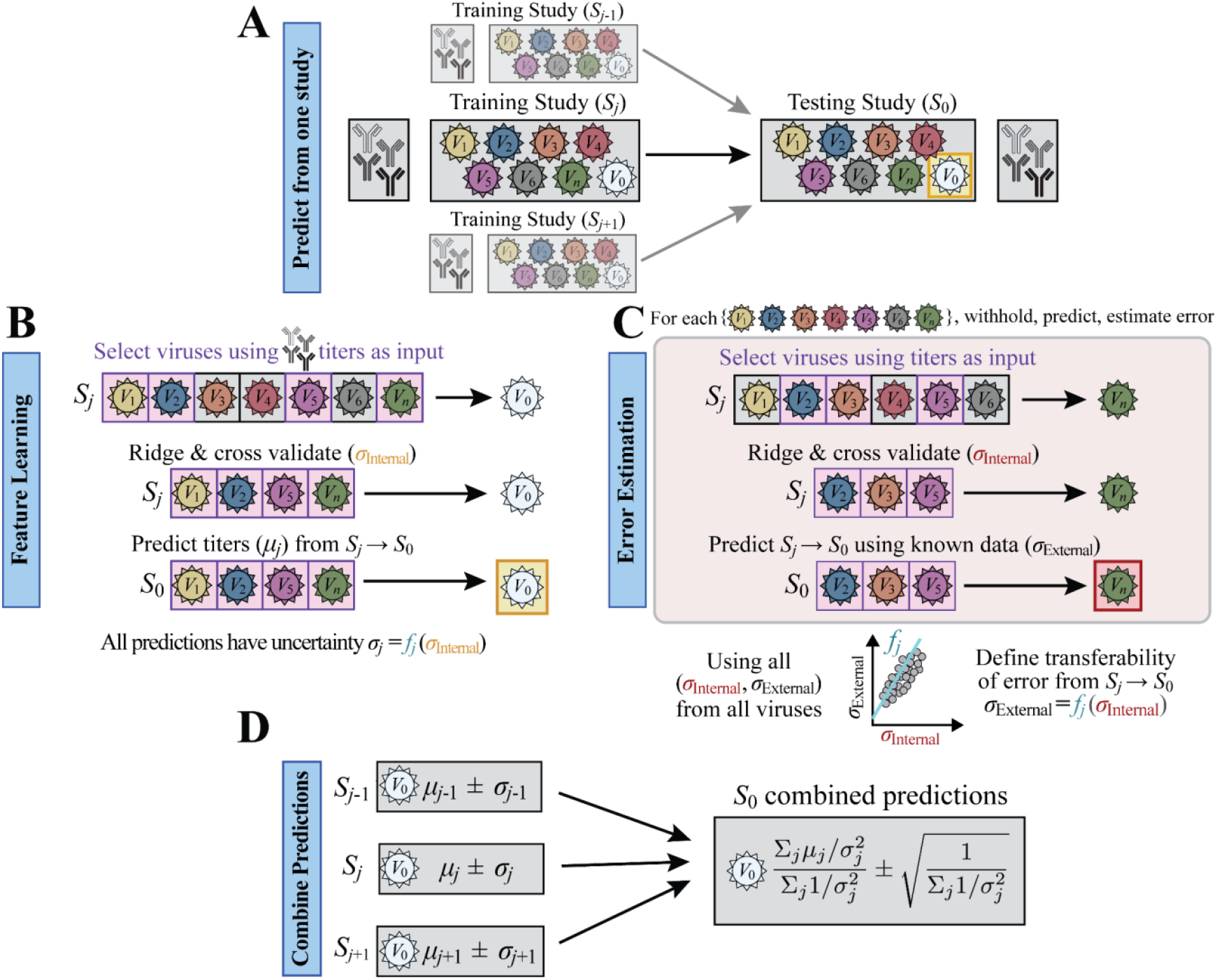
Combining datasets using CAPYBARA to predict new antibody-virus interactions. (A) Given studies (…*S*_*j*-1_, *S*_*j*_, *S*_*j*+1_…) measuring serum HAI against a subset of influenza variants *V*_0_-*V*_*n*_, and study-of-interest *S*_0_ measuring HAI against *V*_1_-*V*_*n*_, CAPYBARA predicts *V*_0_’s measurements in *S*_0_. (B) CAPYBARA first identifies the most predictive variant subsets using Recursive Feature Machines (purple). Ridge regression is applied using those features, training on a subset of data in *S*_*j*_ and cross-validating on the rest (error *σ*_Internal_). This model predicts titer values *μ*_*j*_ from *S*_*j*_ → *S*_0_ without uncertainty. (C) To estimate cross-study prediction error, every other variant is withheld and predicted from *S*_*j*_ → *S*_0_ to determine the internal (*σ*_Internal_) and cross-study (*σ*_External_) error. Combining the errors from every overlapping variant yields the transferability function *f*_*j*_ that is applied to *V*_0_’s *σ*_Internal_ from Panel B to estimate the uncertainty *σ*_*j*_ in *S*_*j*_. (D) Predictions from all studies are combined through a Bayesian approach to yield a consensus prediction for the study-of-interest (*S*_0_).

A key innovation from previous methods is that this model combines state-of-the-art feature selection^24^ and error estimation techniques^25^ while leveraging ridge regression for greater interpretability. CAPYBARA identifies the most predictive virus variants using a Recursive Feature Machine (RFM) that equips general machine learning models with the ability to learn features from data.^24^ The resulting features are used to train an additional ridge regression algorithm, with error estimation performed by predicting all overlapping data between each pair of studies.^25^ The final model predictions are combined using Bayesian weighing. We demonstrate that this approach is robust and generalizable by applying CAPYBARA to 25 influenza HAI datasets, which to our knowledge comprises one of the largest compilations of influenza serology studies measuring multiple variants.

In the context of influenza immunity, CAPYBARA addresses two essential questions: First, how accurately can we leverage prior studies to predict future antibody inhibition data? Second, how few measurements are needed in order to extrapolate all antibody-virus interactions for any set of variants? By predicting these interactions, CAPYBARA can not only expedite future experiments but also explicitly and unbiasedly quantify how differences in study populations, experimental conditions, and virus panels impact the magnitude, breadth, and resolution of the immune response.

## Results

### Overview of the Algorithm

The CAPYBARA algorithm predicts the HAI titers of multiple sera against a withheld or unmeasured variant-of-interest *V*_0_ in study-of-interest *S*_0_. As input, we assume that HAI titers from other variants *V*_1_, *V*_2_, *V*_3_… were measured for these same sera, and that other studies *S*_1_, *S*_2_… also measured HAI for *V*_0_ and a subset of other variants (**Fig 1A**).

The algorithm proceeds as follows: 1) In every other study *S*_*j*_∈{*S*_1_, *S*_2_…}, identify the most predictive subset of variants (features) that predict HAI titers for *V*_0_ (**Fig 1B**). 2) Train a model in *S*_*j*_ to predict each subject’s titer (*μ*_*j*_) for *V*_0_ (**Fig 1B**). 3) Repeat step 2 on all other variants *V*_1_, *V*_2_, *V*_3_… whose values are known, so that within-study and cross-study error can be computed. This determines how the error relationship when predicting from *S*_*j*_ to *S*_0_ (**Fig 1C**), which is then applied to determine the uncertainty *σ*_*j*_ for *V*_0_ predictions in *S*_*j*_. 4) Combine predictions from all studies to estimate the HAI titer±error for each subject (**Fig 1D, Methods**). Prediction accuracy can only decrease as more datasets are included, although adding a very noisy dataset (*σ*_*j*_→∞) will negligibly change predictions.

To validate how well CAPYBARA predicted unmeasured serum-virus interactions across a compendium of influenza studies, we entirely withheld antibody responses from each variant within 20 vaccine studies and 5 longitudinal infection studies conducted between 1997-2023 (**Table 1**). These studies covered a variety of vaccine types (inactivated, live attenuated), age groups (children and adults), and geographic regions, containing ∼200,000 HAI titers from 3,855 unique subjects (**Table 1, Fig S1**), Given this diversity, it was unclear *a priori* which datasets would be most informative to impute the HAI titers in any other study.

**Table 1.**
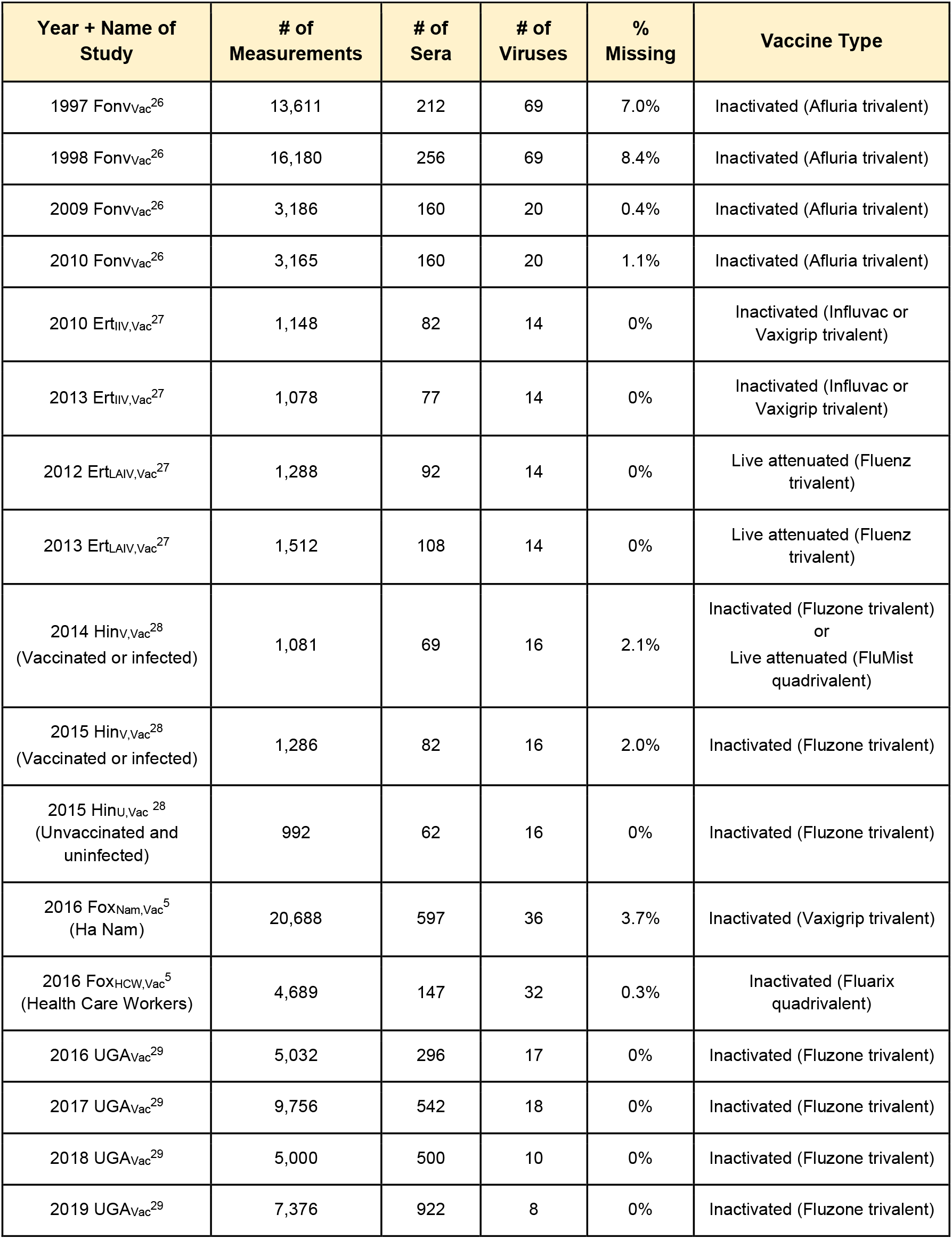
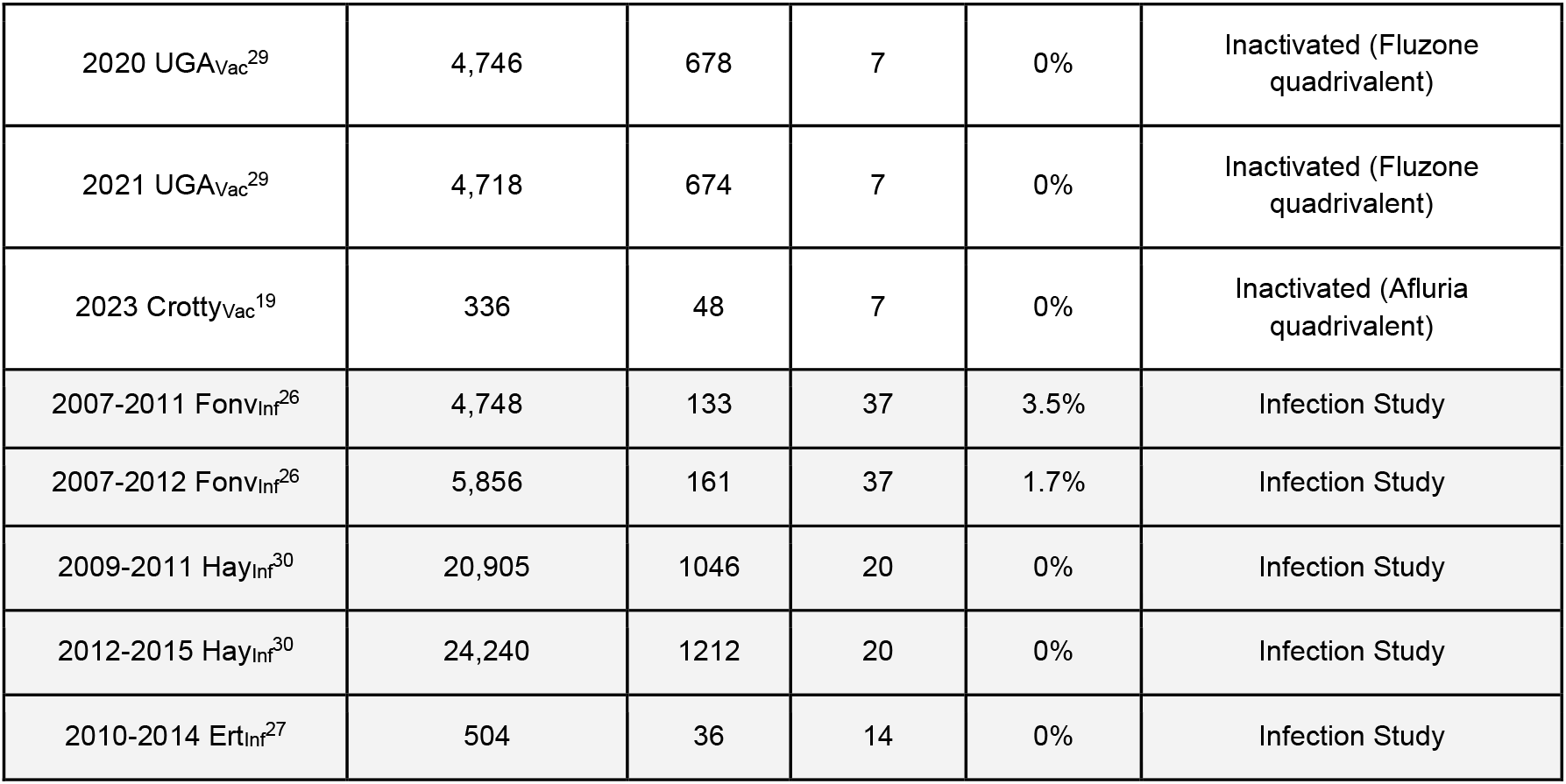
List of large-scale influenza studies used in this analysis. 25 influenza datasets comprising vaccine [Vac, white background] or infection studies [Inf, gray background] used to assess cross-study predictions. The year represents when each study was conducted (*e*.*g*., 2010-2014 implies that samples were collected annually across these 5 years). Sera collected at different time points from the same subject were considered independently. The total number of measurements in each study equals (# of sera)×(# of viruses)-(% Missing) denotes a missing antibody-virus HAI

### Antibody responses are predicted between infection and vaccination studies within experimental noise

To test how well the HAI of new variants could be inferred across diverse biological contexts, we first examined how a longitudinal 6-year infection study (2007-2011 Fonv_Inf_) predicted the overlapping variants in a vaccine study conducted six years later (2017 UGA_Vac_), by which point subsequent infections or vaccinations could have dramatically altered HAI cross-reactivity. In total, *N*=4,336 titers were imputed in the vaccine study with root-mean-squared error (RMSE) *σ*_Actual_=3.1x (where “x” denotes fold-error) between the predicted and measured titers (**Fig 2A**, blue), implying that a measured HAI=20 will typically be predicted as a titer between 20/3.1=6.5 and 20·3.1=62. The model’s estimated error *σ*_Predict_=7.7x represents an upper bound (worst case) error, and although this bound was not tight, it satisfied *σ*_Actual_≲*σ*_Predict_ as expected. In contrast, when we predicted this same 2017 UGA_Vac_ dataset using a vaccine study from one year earlier (2016 UGA_Vac_), we found a smaller prediction error *σ*_Actual_=2.0x and a tighter estimated error *σ*_Predict_=2.3x when predicting these same *N*=4,336 titers (**Fig 2A**, red).

**Figure 2.**
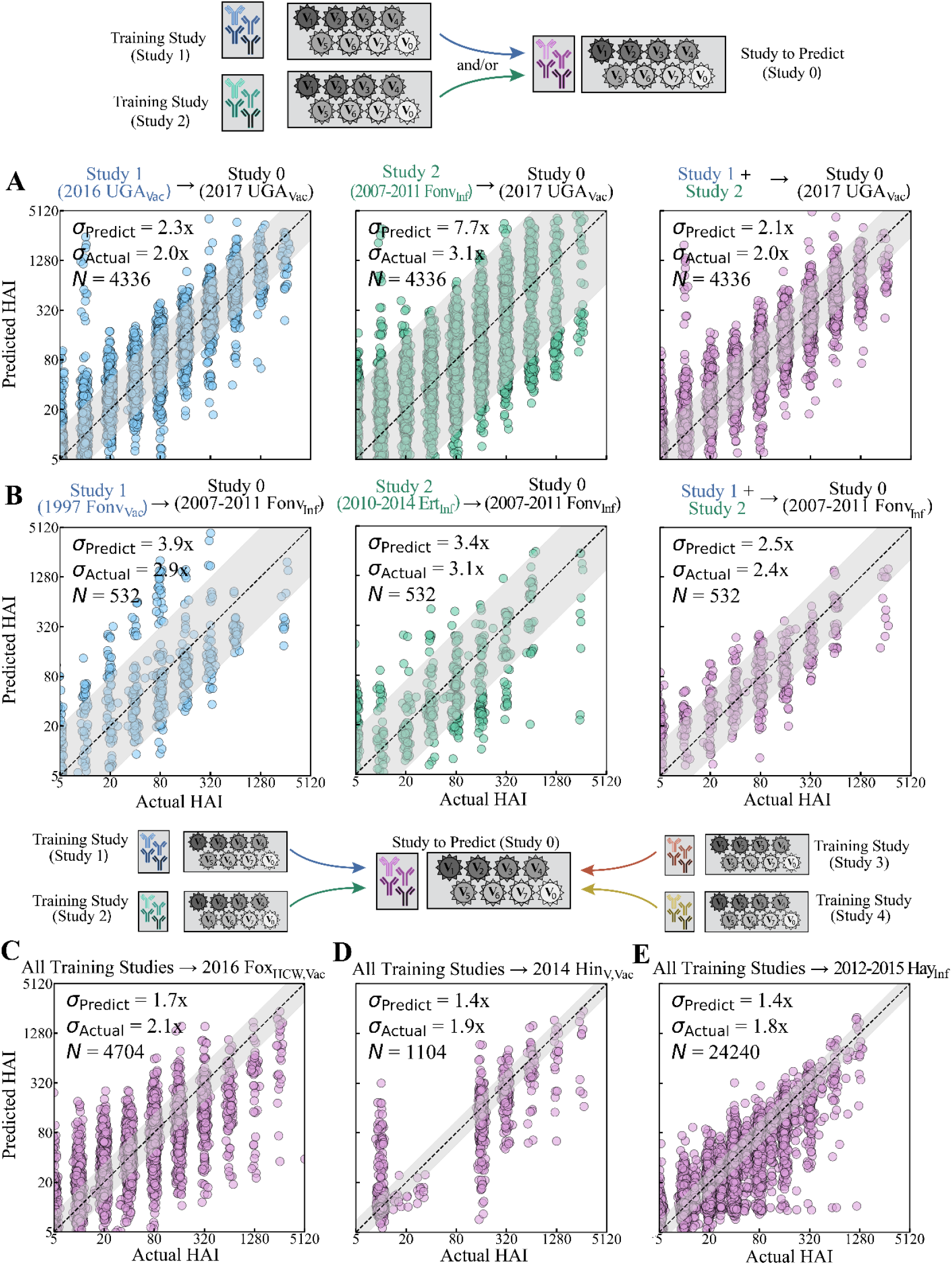
HAI titers across vaccination and infection studies are consistently predicted within experimental noise by combining predictions from all other studies. (A,B) Example predictions trained on an individual dataset (left and middle columns) and the combination of both datasets (right column). Labels above each plot identify the training → testing datasets. (C-E) Predicting three datasets using all other studies in **Table 1**. The estimated fold-error (*σ*_Predict_), measured fold-error (*σ*_Actual_), and the number (*N*) of predicted titers are shown, with the gray diagonal bands representing *σ*_Predict_.

For each serum-virus HAI, the estimated titer and error (*μ*_1_±*σ*_1_ from study 1, *μ*_2_±*σ*_2_ from study 2) were combined using Bayesian statistics, (*μ*_1_/*σ*_1_^2^+*μ*_2_/*σ*_2_^2^)/(1/*σ*_1_^2^+1/*σ*_1_^2^) ± (1/*σ*_1_^2^+1/*σ*_2_^2^)^-1/2^, which places more weight on the more confident prediction with a smaller *σ*_Predict_ (**Methods**). In this case, 2016 UGA_Vac_ was weighed ∼20x more heavily (1/*σ*_1_^2^=0.19 vs 1/*σ*_2_^2^=0.01), as may be expected from its similar study design. The combined predictions remained as good as the predictions from the 2016 UGA_Vac_ study alone (*σ*_Actual_=2.0x, *σ*_Predict_=2.1x), demonstrating that the model is not hampered by adding the poorly predicted infection study (**Fig 2A**, purple).

As another example, we used an infection study (2010-2014 Ert_Inf_) and a vaccine study (1997 Fonv_Vac_) to both individually and jointly predict another infection study (2007-2011 Fonv_Inf_). Interestingly, predictions between the infection→infection study (*σ*_Actual_=3.1x) were very slightly worse than vaccine→infection predictions (*σ*_Actual_=2.9x), even though the infection studies overlapped in time while the vaccine study occurred ten years earlier (**Fig 2B**, blue/green). Combining both studies led to more accurate predictions than either dataset alone (*σ*_Actual_=2.4x) with similarly tight estimated error (*σ*_Predict_=2.5x, **Fig 2B**, purple).

More generally, the predictions from any number of datasets can be combined using error estimation (**Methods**). As representative examples, we used every dataset in **Table 1** to predict HAI titers in an adult health care workers vaccine study (2016 Fox_HCW,Vac_, **Fig 2C**), vaccinated children (2014 Hin_V,Vac_, **Fig 2D**), and an adult infection study (2012-2015 Hay_Inf_, **Fig 2E**; all studies in **Fig S2**). The 10^3^-10^4^ predicted HAIs in each study had low RMSE (*σ*_Actual_=1.8-2.1x, similar to the error of the HAI assay) and comparably low estimated error (*σ*_Predict_=1.4-1.7x), demonstrating that combining datasets can precisely and confidently extrapolate HAI titers for completely unmeasured variants. These results corroborate that studies do not need to be pre-screened, since the framework will identify the most predictive datasets and ignore the poorly predictive ones.

### More training datasets lead to better prediction accuracy

To test generalizability, we compared the prediction error when training on any single dataset vs the combined predictions from all studies (**Fig 3A**). As expected, the individual datasets showed far larger variability (*σ*_Actual_=1.6-10.7x) than the combined predictions (1.7-2.5x) that are always comparable to the most accurate pairwise predictions in each column. Interestingly, prediction accuracy does not need to be symmetric. For example, multiple studies had poor prediction with >6x when trained on 1997 Fonv_Vac_, yet using any training dataset to predict values in 1997 Fonv_Vac_ always led to accuracy <4x.

**Figure 3.**
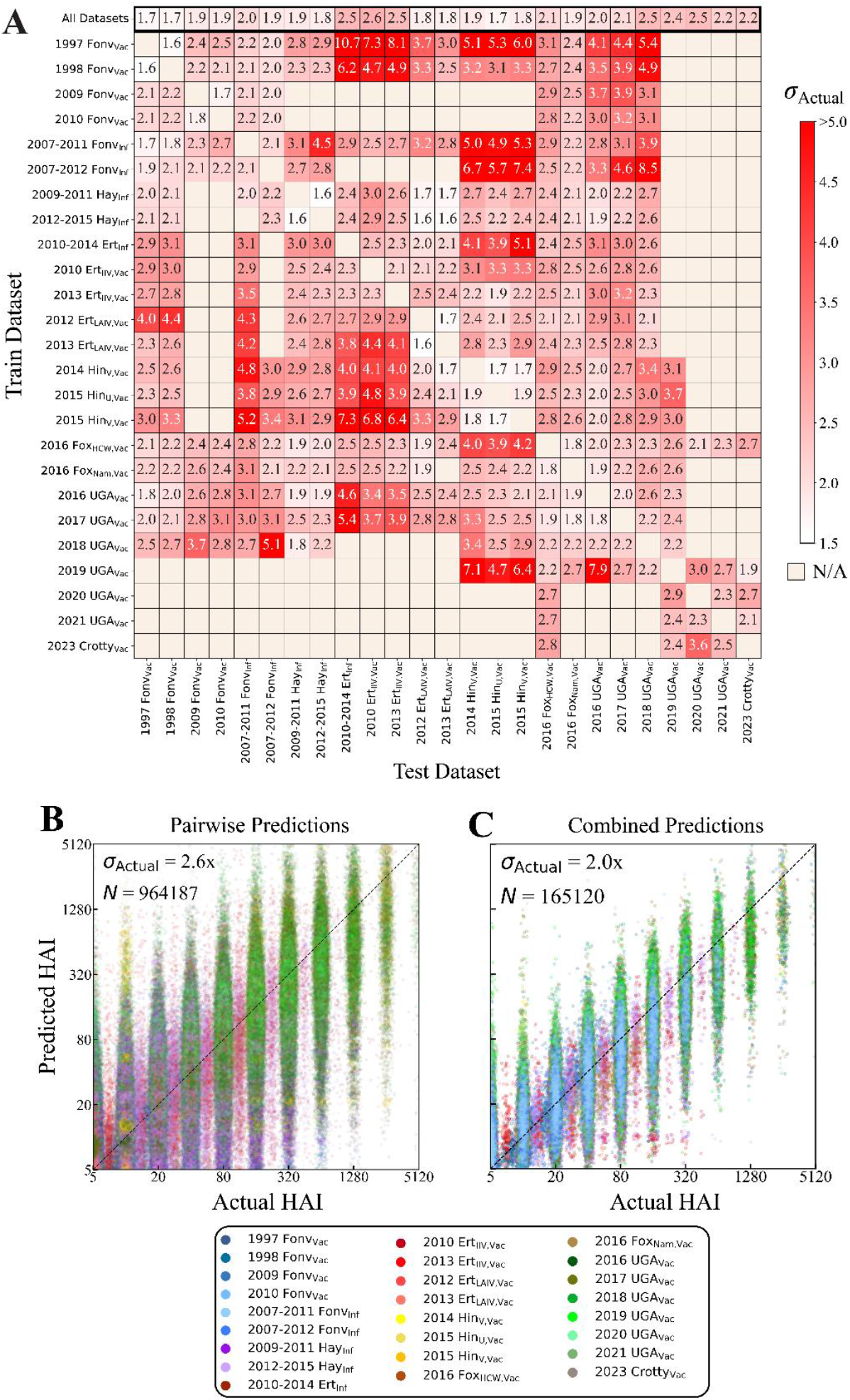
Predicting HAI responses across all studies. (A) Heatmap of the average RMSE (*σ*_Actual_) across all subjects and overlapping variants in a study-of-interest (column). Training is either done using all studies (top row) or using a single study (all other rows). (B-C) All predicted versus measured HAIs when training on (B) a single study or (C) all other studies. The number *N* of predictions is larger for pairwise predictions since the same serum-virus pair is predicted multiple times using different training datasets. The diagonal line *y*=*x* represents perfect predictions.

At the individual-person level, there is a noticeably greater spread in pairwise predictions (*σ*_Actual_=2.6x across all subjects, **Fig 3B**) than in the combined predictions (2.0x, **Fig 3C**), with 14.3% of the former predictions having an error >4x while only 5.3% of the latter predictions have such error (**Fig S3**). Indeed, CAPYBARA does better than averaging the individual predictions from each study by heavily weighing the more reliable, and hence more accurate, predictions (**Fig S4**).

A few general trends can be seen from pairs of studies that poorly predict one another (**Fig 3A**). The two oldest studies (1997/1998 Fonv_Vac_) tend to poorly predict studies from 2010 and beyond. The LAIV studies (2012/2013 Ert_LAIV,Vac_) were sometimes poorly predicted by the more common IIV studies. Beyond these few rules, it was often unclear which studies would poorly predict one another, emphasizing the utility of CAPYBARA to infer such relationships directly from the data.

### Subsetting datasets helps explain prediction dynamics

Since age is well known to affect the antibody response, we assessed how well children (age≤18) can predict adult responses (age>18) and vice versa. Datasets were categorized as containing children only, adults only, or a combination of both (**Fig S1**). HAI titers from studies in each category were exclusively predicted using models from either the same or a different category (**Fig 4A**). As expected, the best predictions came from models trained within the same category. For example, children’s titers were better predicted by children data (*σ*_Actual_=1.7x) than by adult data (*σ*_Actual_=2.4x; *p*<0.05, two-sided permutation test). Studies containing both children and adults represented an intermediate phenotype, which was itself best predicted by studies containing titers from children and adults. Despite most of these age effects being significant, the overall effect of age was small, where even purposefully mismatching datasets (predicting children→adults or adults→children) led to median error <2.5x.

**Figure 4.**
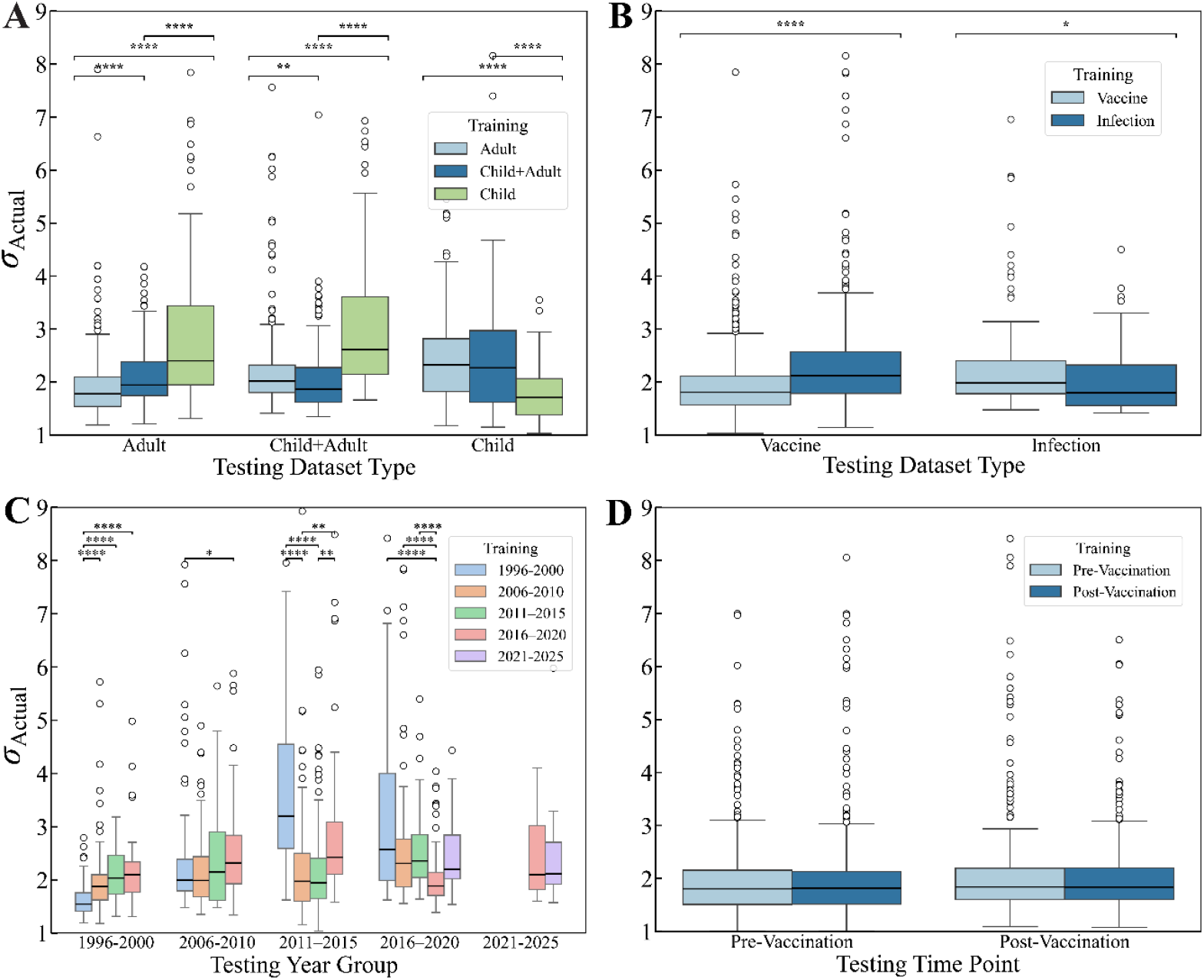
Training on similar datasets marginally improves prediction accuracy. Cross-study RMSE (*σ*_Actual_) when training and predicting between datasets based on (A) the age groups adult-only, children-only, or mixed (child + adult); (B) vaccination or infection studies; (C) datasets grouped in 5-year intervals based on their median year; or (D) pre-vaccination (Day 0) vs post-vaccination (∼1 month) data. Each box plot shows the distribution of errors for all possible withheld variants. The horizontal line denotes the median, boxes show the interquartile range, and whiskers extend to 1.5 times the interquartile range. Circles denote outliers. Statistical significance was assessed using two-sided permutation tests with Benjamini–Hochberg correction for multiple testing. Asterisks denote adjusted *p*-values: **** = *p*<0.0001, *** = *p*<0.001, ** = *p*<0.01, * = *p*<0.05.

We next split datasets by study type (vaccine versus infection). As before, there was a small but significant improvement in prediction accuracy when the same type of dataset was used for training (**Fig 4B**). For example, predicting from infection→infection studies (*σ*_Actual_=1.7x) was more accurate than vaccination→infection (*σ*_Actual_=2.0x; *p*<0.05, two-sided permutation test), although predictions in either case were surprisingly accurate, with similar results when predicting vaccination responses.

The worst prediction accuracy was seen when splitting datasets by their year of study and using old datasets to predict responses >10 years into the future (**Fig 4C**). Studies were binned in five year increments, with studies conducted over multiple years represented by their median year. Training on studies from the same bin either led to the best predictions or to comparable predictions with the best bin (median *σ*_Actual_ 1.6–2.1x). Accuracy dropped, often significantly, when using older datasets to predict more recent ones. In particular, training the oldest 1996– 2000 datasets led to poor predictions and large variation on 2011–2015 (*σ*_Actual_ = 3.2x; *p*<0.05, two-sided permutation test) or 2016–2020 data (*σ*_Actual_ = 2.6x; *p*<0.05, two-sided permutation test), although predictions going backwards in time by >10 years tended to be more accurate.

Lastly, we examined how accurately pre-vaccination titers predicted the peak post-vaccination titers (21-43 days post-vaccination) across vaccine studies. Surprisingly, we observed nearly identical prediction accuracies (median within ∼0.02x of each other; p=1.0, two-sided permutation test), suggesting that the HAI cross-reactivity across variants holds over time, with most variants increasing in tandem post-vaccination.

### Identifying universal relations between influenza variants

To demonstrate how future studies can leverage CAPYBARA to measure a few variants and infer the response from others, we sought universal relations that could be applied to a new study without requiring dataset reweighing through CAPYBARA. To that end, we used RFM to denote which variant features were the most important when predicting each of the 112 variants across these studies (**Fig 5A, Fig S5**; red represents greater importance).

**Figure 5.**
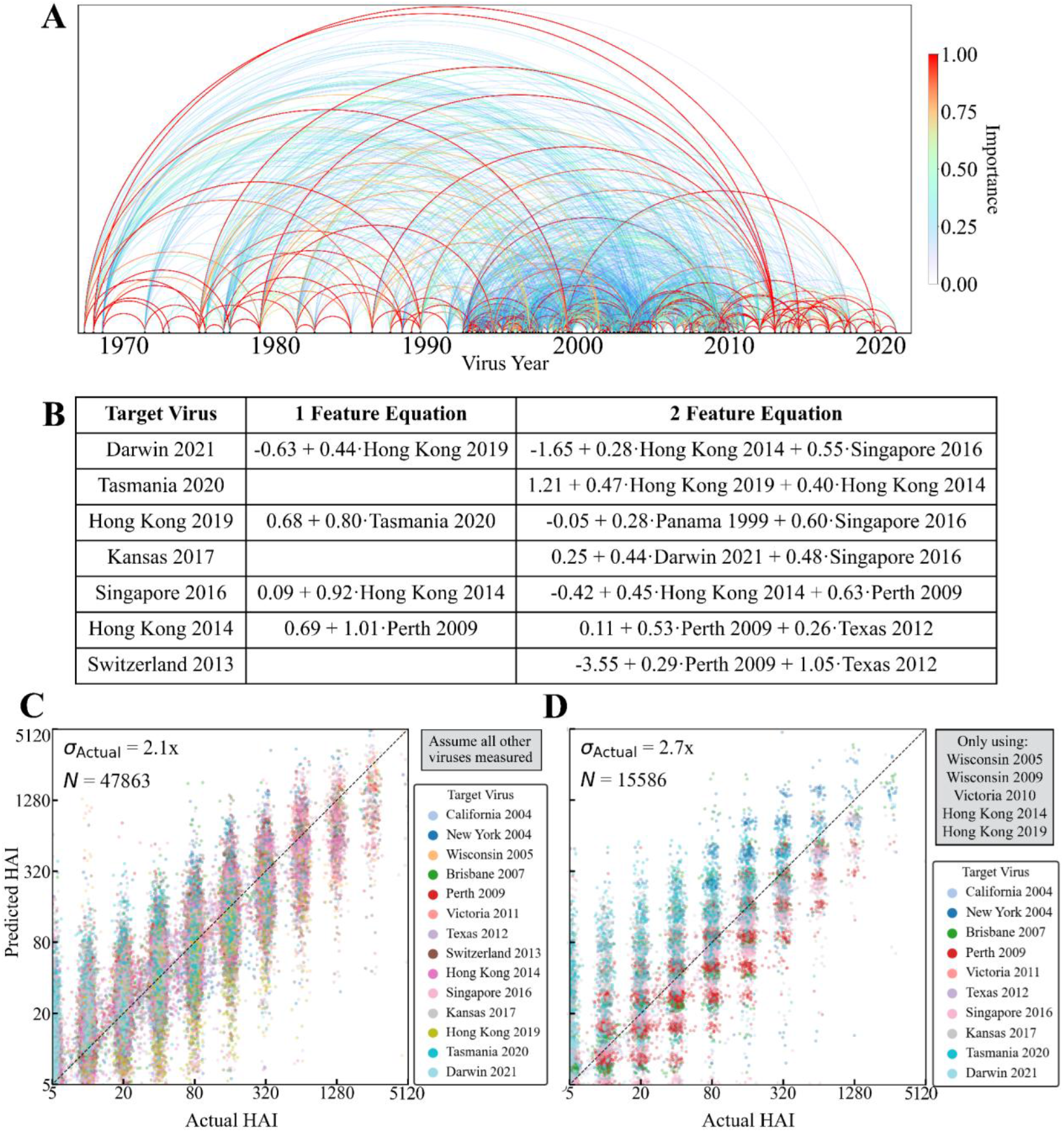
A global dictionary of influenza variant importance. (A) Rainbow diagram of feature importance between any pair of variants (connections are bidirectional). (**B**) Examples of universal HAI titer equations for multiple influenza vaccine strains, using titers from one variant (when possible) or two variants. Each virus name stands for the log_2_(HAI/5) of its titer. See the **Supporting Information** for all relations using ≤5 variants. (**C**) Measured versus predicted HAI titers for all vaccine strains in each study. Predictions were averaged from all other studies that measured the necessary variants. (**D**) Example using a small subset of five variants to predict ten other vaccine strains.

While distant strains circulating more than 10 years apart could be important (average feature importance=0.3), the most important variant pairs tended to circulate less than a decade apart (average feature importance=0.5, **Fig S5**). However, feature importance could only be determined when two viruses were measured in at least two studies, so that the more frequently selected vaccine strains tend to have far better coverage than non-vaccinate strains. For example, the 1968 pandemic strain Hong Kong 1968 was often measured, and it exhibited strong feature importance of ≈1 against viruses circulating as late as Hong Kong 2014.

Each variant-of-interest *V*_0_ in study *S*_0_ is predicted by every other study with at least three overlapping strains, leading to multiple potential distinct HAI relations. While 53.1% of relations required 1-2 variants, 20.3% of equations required 4 or more variants (all relations shown in **Supporting Information**). Since vaccine strains were frequently measured, many relations exclusively use these strains (examples in **Fig 5B**).

To evaluate the accuracy of these relations, each vaccine strain’s HAI titers were individually withheld from a study-of-interest and derived by averaging the relations from all other studies. To make these results as readily generalizable as possible, predictions were not weighted by their estimated accuracy as in the sections above, but instead averaged equally across all studies. The resulting predictions showed an RMSE of 2.1x, demonstrating that directly averaging these relations leads to remarkably accurate predictions with error comparable to the ≈2-fold error of the HAI assay, provided that each study measures all of the necessary variants to apply these relationships (**Fig 5C**).

To further expedite future studies, we assessed whether measuring a smaller set of only five influenza variants (comprising four vaccine strains and one non-vaccine strain) could predict ten other vaccine strains as well as two non-vaccine strains (**Fig 5D, Supporting Information**). This reduced set of variants only had a slightly larger RMSE of 2.7x, demonstrating that cross-study relationships can increase the amount of data generated by a few experiments, and that prediction accuracy should increase as more datasets are measured, or by applying CAPYBARA to heavily weigh the most accurate studies.

## Discussion

Here, we developed CAPYBARA, a general algorithm that combines feature learning, model generation, and error estimation to predict unmeasured interactions based on existing datasets. As a case study, CAPYBARA was applied to identify universal patterns in serum-virus cross-reactivity and predict each serum’s HAI against variants that were entirely withheld from a study.

While factors such as age ^7-10,31^ or exposure history^1,3,5,6^ are known to affect the antibody response, it is unclear how these impact serum cross-react across influenza variants. To that end, CAPYBARA quantifies how accurately the local relationships in one dataset translate into another dataset using all non-withheld data, testing this approach across 25 different influenza studies.

A key piece of this approach was to use error estimation to quantify the transferability between datasets, since some studies poorly predicted a dataset-of-interest with error>4x, while others were accurate within the ≈2x intrinsic error of the HAI assay. The combined predictions based on Bayesian weighing consistently favored the most informative datasets, leading to 1.7-2.5x prediction error across all studies.

The algorithm did require any prior information about subject demographics, study design (vaccination vs infection, time points measured), or exposure history. Instead, CAPYBARA unbiasedly used overlapping variant HAI titers to determine how well one dataset can predict another. With these results, we retrospectively examined how these various factors affected the antibody response.

Subject age had a small but significant effect, suggesting that cross-reactivity changes from childhood (age≤18) into adulthood (age>18). Children predicted other children’s responses better than adults, while adults predicted other adult responses better than children, with mixed datasets containing both children and adults falling in the middle. The year a study was conducted also had a significant effect, with studies within a 10 year window exhibiting 1.6x-2.4x error while studies done further apart in time had 2.0x-3.2x error. Vaccination and infection studies similarly predicted their own category better than the other category. Surprisingly, within vaccine studies, the pre-vaccination (day 0) and peak response (day 21-40) time points predicted one another with comparable accuracy, suggesting that pre- and post-vaccination cross-reactivity resemble one another. This could arise if all variant HAIs increase by a similar amount post-vaccination, or if post-vaccination responses are relatively weak, both of which held true across these datasets and were previously reported.^32^

One limitation of this approach is that a variant’s HAI titers can only be predicted in a dataset-of-interest if that variant has been measured in at least one other study. Thus, this method is not equipped to predict the HAI of new variants, although a variant measured in one dataset can be predicted in all other studies. As datasets measuring more variants are added, the number of predictions in each study grows combinatorially.

As such, CAPYBARA lays the foundation to design more efficient experiments that leverage existing studies. It further provides a quantitative foundation to determine the minimum number of variants that should be measured to infer the HAIs from multiple variants of interest. To facilitate such use, we also provide the average cross-reactivity relations between all H3N2 influenza variants examined in this work (**Supplementary Information**). These relations can be immediately applied to a new study, or they can be further augmented with CAPYBARA that will derive new dataset-specific relations weighed by dataset similarity.

## Methods

### Overview of the datasets

We analyzed a collection of 25 influenza vaccine and infection studies spanning 1997–2023 (**Table 1**). If one participant had multiple sera (*e*.*g*., pre-vaccination and post-vaccination), the two were analyzed independently. Predictions were carried out between two datasets if they measured HAI against at least three of the same H3N2 variants, since this ensures that there are enough features for cross-study prediction.

### Analyzing HAI Titers

All studies used hemagglutination inhibition, which measures the highest dilution of serum at which hemagglutination is inhibited. A larger HAI titer will reflect a more potent serum, but it may also reflect differences in virus passaging (egg-vs cell-grown) or study design (incubation conditions, type or batch of red blood cells). Missing HAI data, comprising 2.1% of all measurements, were imputed using the row–column mean, and these imputed values were both predicted and also used to predict other titers.

As in prior analyses, titers were transformed to log_2_(HAI/5), which reduces the bias toward large titers.^14,25^ All prediction errors are shown in unlogged units so that they can be compared to the measured HAI titers. More precisely, the root-mean-squared error (*σ*_Actual_) of the logged titers is exponentiated by 2 to get the unlogged error (*i*.*e*., *σ*_Actual_=1.0 for log_2_ titers corresponds to an error of *σ*_Predict_=2^1.0^=2-fold, with “fold” or “x” indicating an un-logged number). Prior work has shown that the HAI has an inherent 2-fold error on average,^19^ and hence predictions with ≈2-fold error are as accurate as possible.

### Overview of CAPYBARA

We first outline the four main steps of the algorithm and then describe each in detail:

**Step 1: Feature Learning** (**Fig 1B**): For each external study *S*_*j*_ that measured the target virus *V*_0_, a Recursive Feature Machine^24^ identifies a small subset of variants that best predict *V*_0_.

**Step 2: Model Training** (**Fig 1B**): Ridge regression is applied to a subset of sera within *S*_*j*_, using the selected variants as inputs and *V*_0_ as the output. The internal root-mean-square error *σ*_Internal_(*V*_0_) is computed on the withheld sera.

**Step 3: Cross-Study Error Calibration** (**Fig 1C**): To extrapolate the regression relation from *S*_*j*_ (where *σ*_Internal_ is known) to the new dataset *S*_0_, CAPYBARA withholds each variant *V*_*k*_≠*V*_0_ and measures how its internal error in *S*_*j*_ maps to its external error *σ*_External_(*V*_*k*_) in *S*_0_. A piecewise linear function is then fit to the (*σ*_Internal_, *σ*_External_) pairs for all *V*_*k*_, and this function is applied to *σ*_Internal_(*V*_0_) to estimate the error in *S*_0_, denoted by *σ*_Predict_(*V*_0_).

**Step 4: Combined Predictions** (**Fig 1D**): When multiple studies can predict a virus *V*_0_ in *S*_0_, their predictions are combined using Bayesian weighting, *i*.*e*., weighting each prediction inversely by its squared predicted error, (1/*σ*_Predict_)^2^. This yields a single predicted HAI titer and a calibrated uncertainty estimate for that titer.

### Step 1: Using Recursive Feature Machines to identify the most predictive features

A Recursive Feature Machine (RFM) is a supervised machine learning model that incorporates feature learning into general non-parametric models through the Average Gradient Outer Product (AGOP).^24^ Unlike prior methods that used brute force (randomly selecting five variants *V*_1_-*V*_5_ to predict a target virus *V*_0_, assessing that selection using cross-validation), RFM gives the feature importance of all variants so that the top candidates can be used to predict *V*_0_. This leads to more efficiently identifying the predictive features, is not restricted to a pre-imposed number of features (*e*.*g*., always requiring five), and yields better predictions than a random search through a subset of possibilities (**Fig S6A**).

Given any differentiable predictor, *f*: ℝ^*d*^→ℝ trained on *n* data points *x*^(1)^, …, *x*^(*n*)^∈ ℝ^*d*^, the AGOP operator *G*(*f*) is the covariance matrix of the input-output gradients of the predictor over the training data,

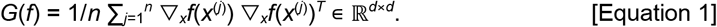

This covariance captures the most predictive directions in its top eigenvectors, and the most important coordinates on its diagonal. RFMs with kernel machines proceeded by obtaining an initial estimate of the target function using a standard kernel machine without feature learning. Given this initial estimate of the predictor, the AGOP of the predictor was computed on the training data, after which the inner product function was updated using the AGOP. RFM then recursed this procedure beginning with the transformed data. Formally, the algorithm proceeded as follows.

#### Algorithm 1: Recursive Feature Machine (RFM)

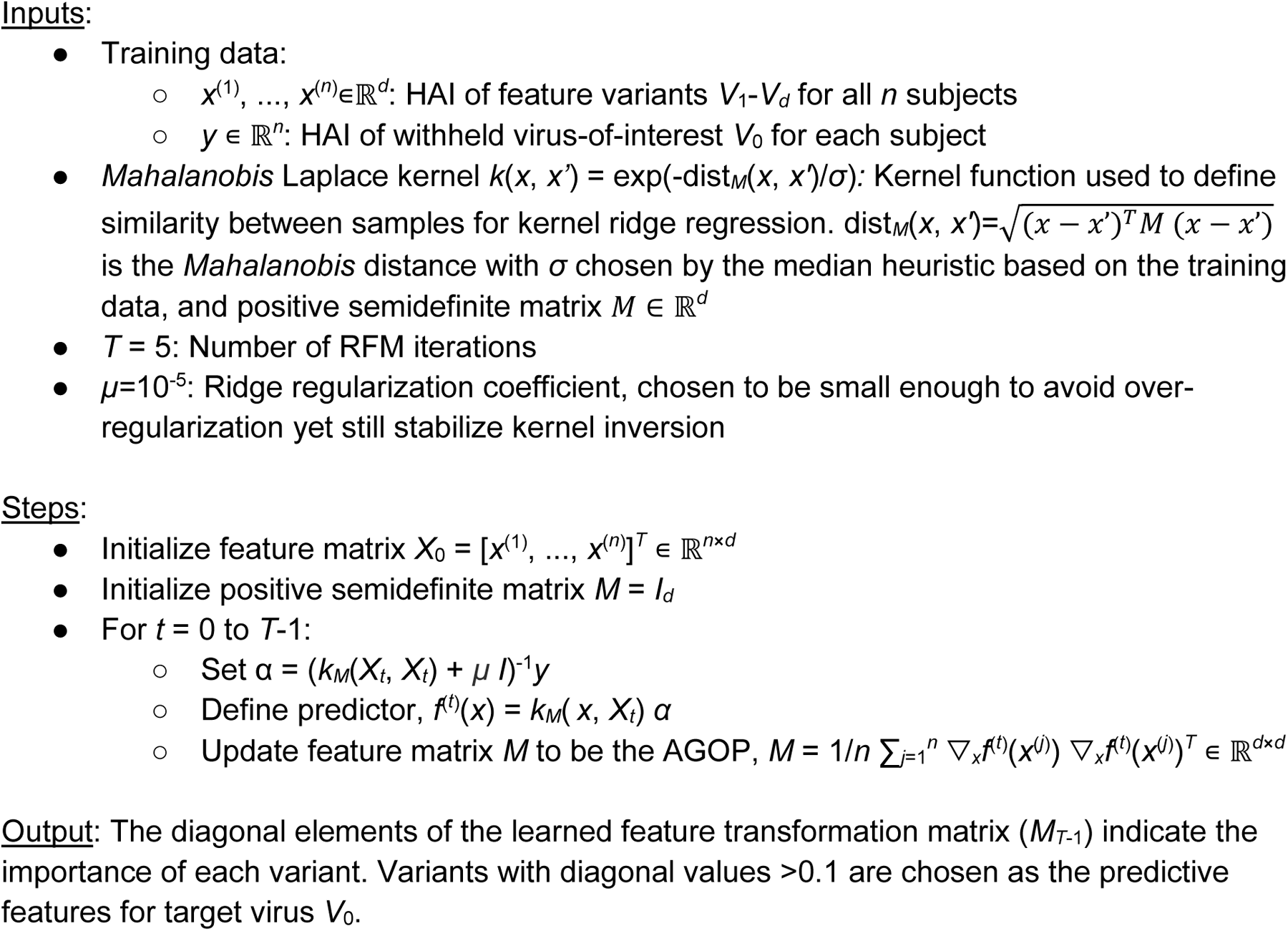

### Step 2: Model training and internal error

Within each external dataset, ridge regression models were trained on selected features identified by RFM. Note that RFM importance was not considered during ridge regression, since in the case of multiple degenerate but highly important features, only a single feature should be selected. Hyperparameters (ridge regularization strength, kernel bandwidth, diagonal thresholds) were optimized via internal cross-validation (80% training, 20% validation splits), but were found to minimally vary (**Fig S6C**).

Following ridge regression, each variant feature with ridge coefficient >0.2 (in absolute value) was retained. When deriving universal cross-reactivity relations, if two studies predicted a target virus *V*_0_ using the same variants as features, the ridge coefficients were averaged for each of the viruses in their equation.

### Step 3: Cross-study error calibration

Following prior work,^25^ to calibrate how accurately the model trained on study *S*_*j*_ applied to study *S*_0_, every possible virus *V*_*k*_≠*V*_0_ was withheld one-by-one (in addition to excluding *V*_0_) from both the training and testing datasets. 80% of sera in the training set were used to fit a ridge regression model in the training dataset, with the remaining sera used to compute the internal error *σ*_Internal_(*V*_*k*_). All sera in the testing dataset were used to compute *σ*_External_(*V*_*k*_). Performing this for all *V*_*k*_ resulted in multiple points (*σ*_Internal_, *σ*_External_) that mapped the transferability of error between the two studies.

These paired internal–external errors were fit using a total-least-squares (orthogonal-distance) line, *σ*_External_ = *α σ*_Internal_ + *β*. To account for the uncertainty of this fit (*i*.*e*., highly scattered points with a poor best-fit line are more uncertain), we added to *σ*_External_ the root-mean-square vertical distance of each point from the fitted line, *δ=*[1/*m*∑_*k*=1_ ^*m*^(*α σ*_Internal_ (*V*_0_) + *β* - *σ* (*V*_External_))^2^]^1/2^. Lastly, the external error was forced to always be at least as large as the internal error. Altogether, the estimated error when predicting variant *V*_0_ in *S*_0_ is given by

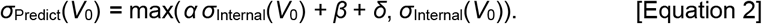

### Step 4: Combining predictions from multiple datasets

When multiple studies *S*_1_, *S*_2_… predicted the HAI titers of virus *V*_0_ in *S*_0_, each subject had predictions *μ*_1_±*σ*_1_, *μ*_2_±*σ*_2_… (where *σ* is a shorthand for *σ*_Predict_). Predictions were combined using Bayesian weighting that is inversely proportional to predicted error squared, namely,

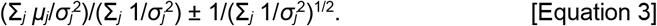

More confident predictions (smaller *σ*_*j*_) are weighted more heavily, while highly inaccurate predictions (*σ*_*j*_→∞) have little-to-no influence. As a result, all datasets can be included, and the algorithm will unbiasedly determine the most accurate predictions and use their values more heavily.

### Software and computational resources

Analyses were implemented in Python using standard scientific libraries (NumPy, SciPy, scikit-learn). Code is available through the accompanying GitHub repository (https://github.com/TalEinav/CAPYBARA).

## Data Availability

Code and data is available through the accompanying GitHub repository

https://github.com/TalEinav/CAPYBARA

## Acknowledgements

We especially thank the experimental groups who shared their data, and we hope this paper will inspire other groups to integrate their datasets for everyone’s benefit. We always welcome pointers to new datasets. We further acknowledge Adit Radha and Mikhail Belkin for useful discussions. This research was supported by LJI & Kyowa Kirin, Inc. (KKNA - Kyowa Kirin North America), and the Bodman family (TE).

## Supplementary Figures

**Figure S1.**
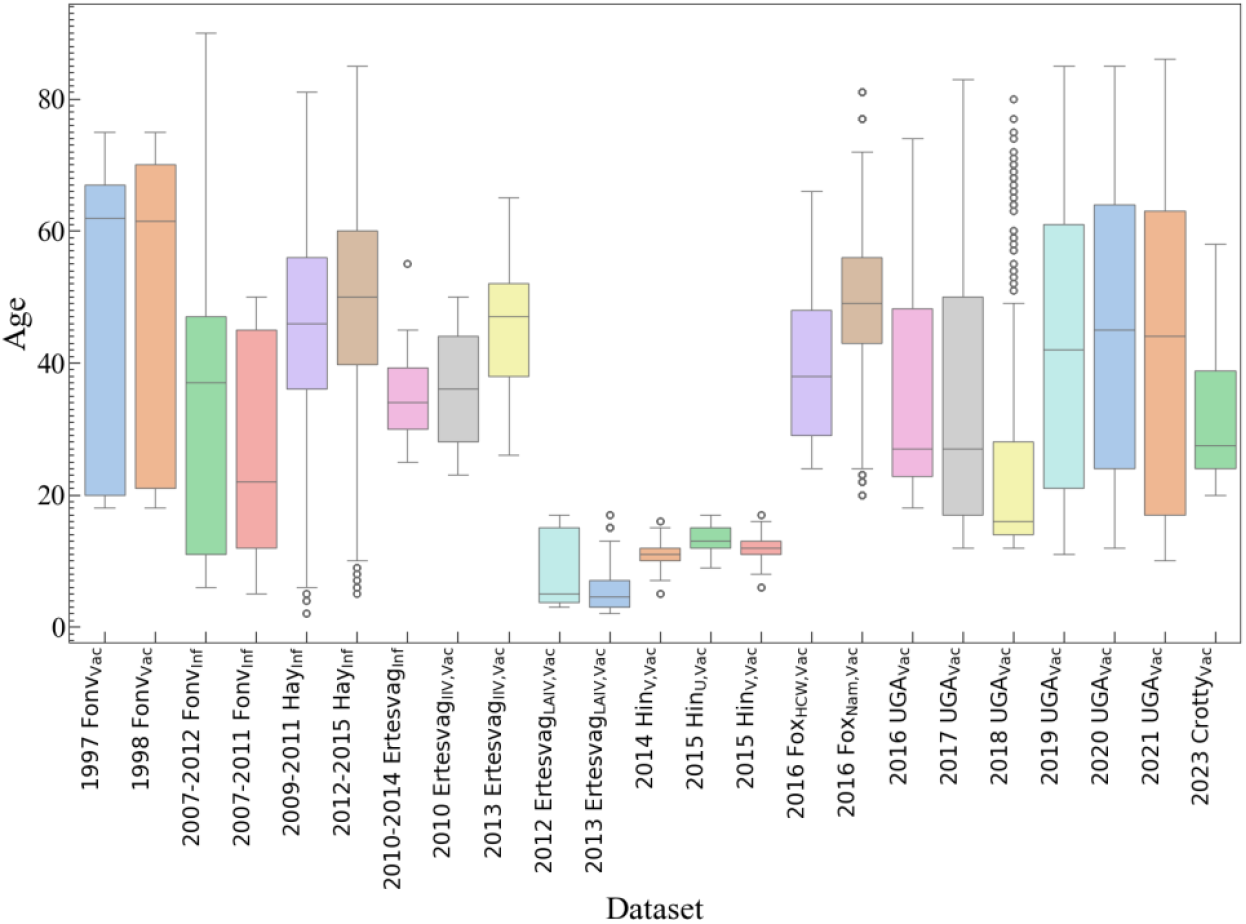
Subject age distributions across datasets. Datasets are ordered chronologically and by study group.

**Figure S2.**
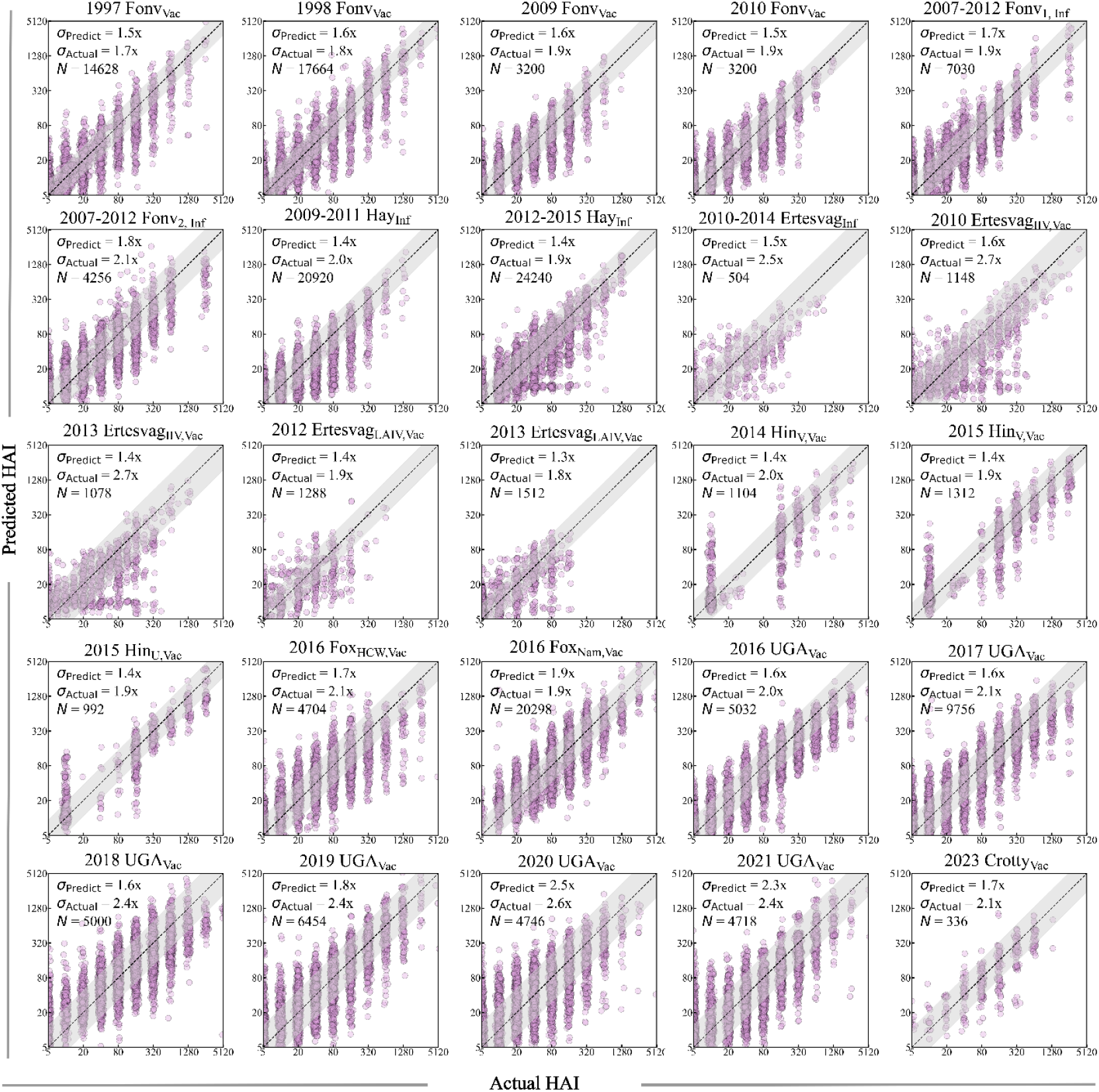
Prediction accuracy using all studies is consistently comparable to experimental noise. Every other study in Table 1 is used to predict HAI titers for all variants in the study-of-interest (shown by the plot label).

**Figure S3.**
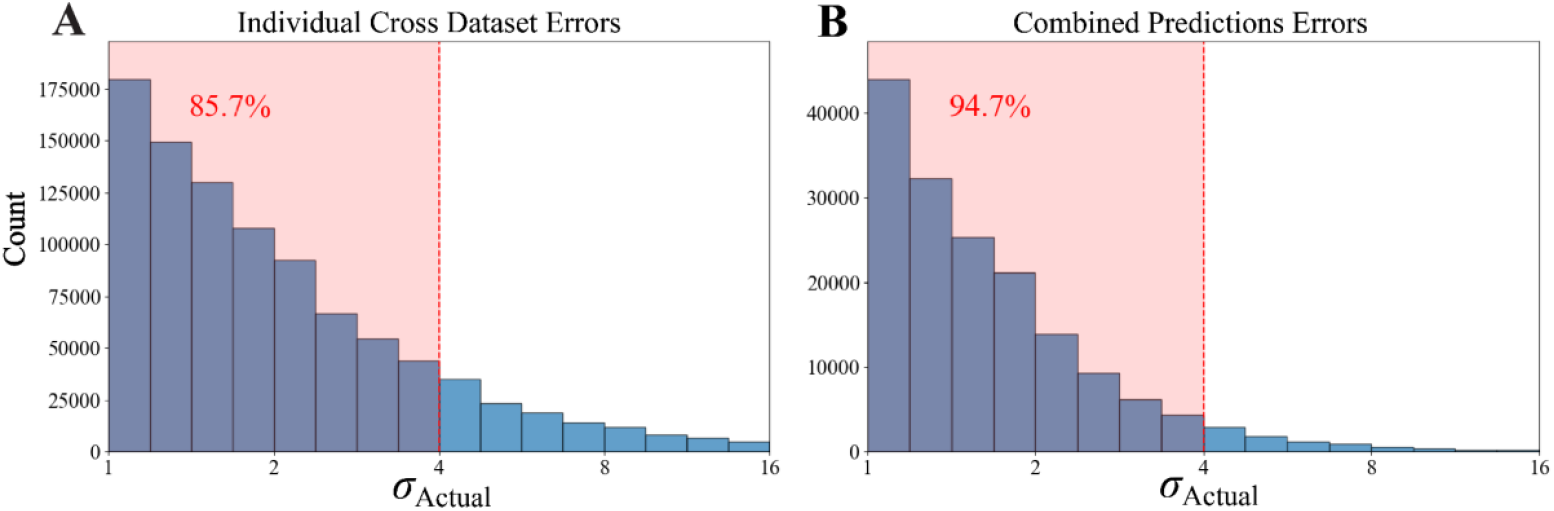
Distribution of errors for individual and combined predictions. Fold-error (*σ*_Actual_) of predictions for every subject and virus using (A) each dataset to make a separate prediction and (B) all datasets to make combined predictions. Red shading marks the region of ≤4x error, and the annotated percentages indicate the fraction of predictions that fall within this threshold.

**Figure S4.**
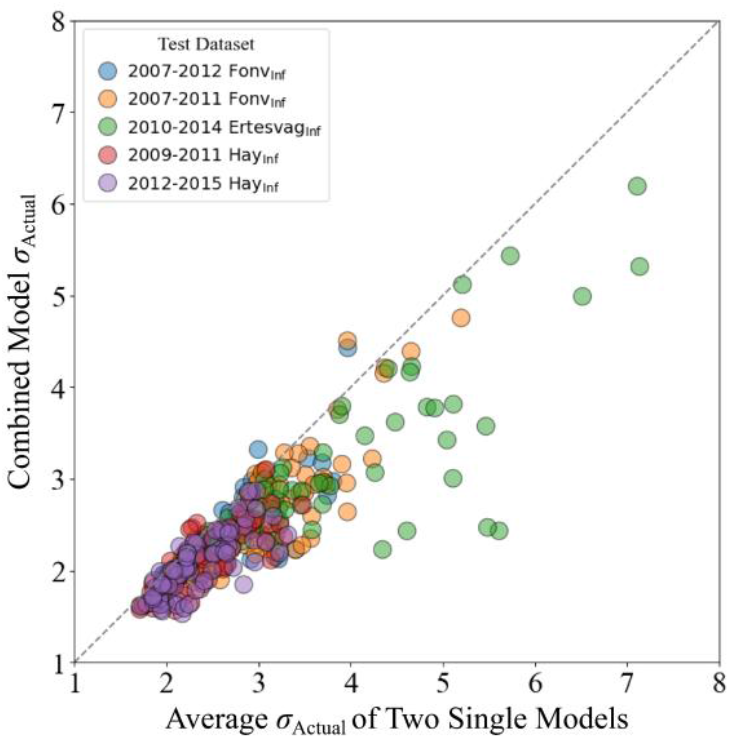
Combined predictions outperform averaged predictions from individual studies. Prediction errors (*σ*_Actual_) for all viruses in all infection studies were computed using two other datasets for training. These two datasets either independently predicted each virus, and their resulting predictions were averaged [*x*-axis] or CAPYBARA was used to combine these predictions by more heavily weighing the dataset that was more similar to the target infection study [*y*-axis]. Points below the diagonal indicate improved performance with the combined model.

**Figure S5.**
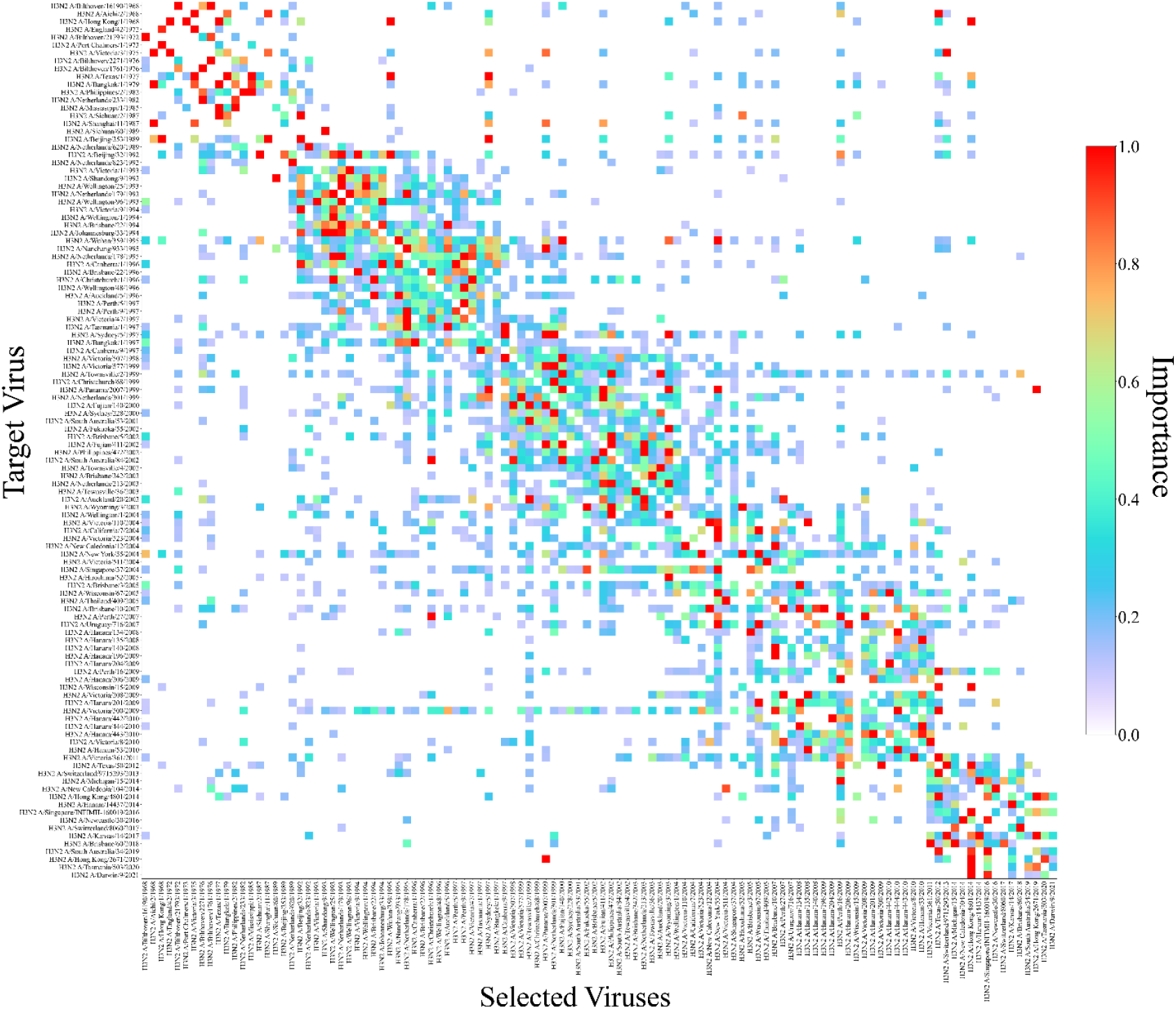
Feature importance via RFM. The importance of each virus feature (column) when predicting a target virus (*V*_0_, row). Feature importance is quantified within a single study. Only viruses with feature importance≥0.1 shown, as these viruses are subsequently used in ridge regression when predicting the target virus. Any virus not picked is shown in white.

**Figure S6.**
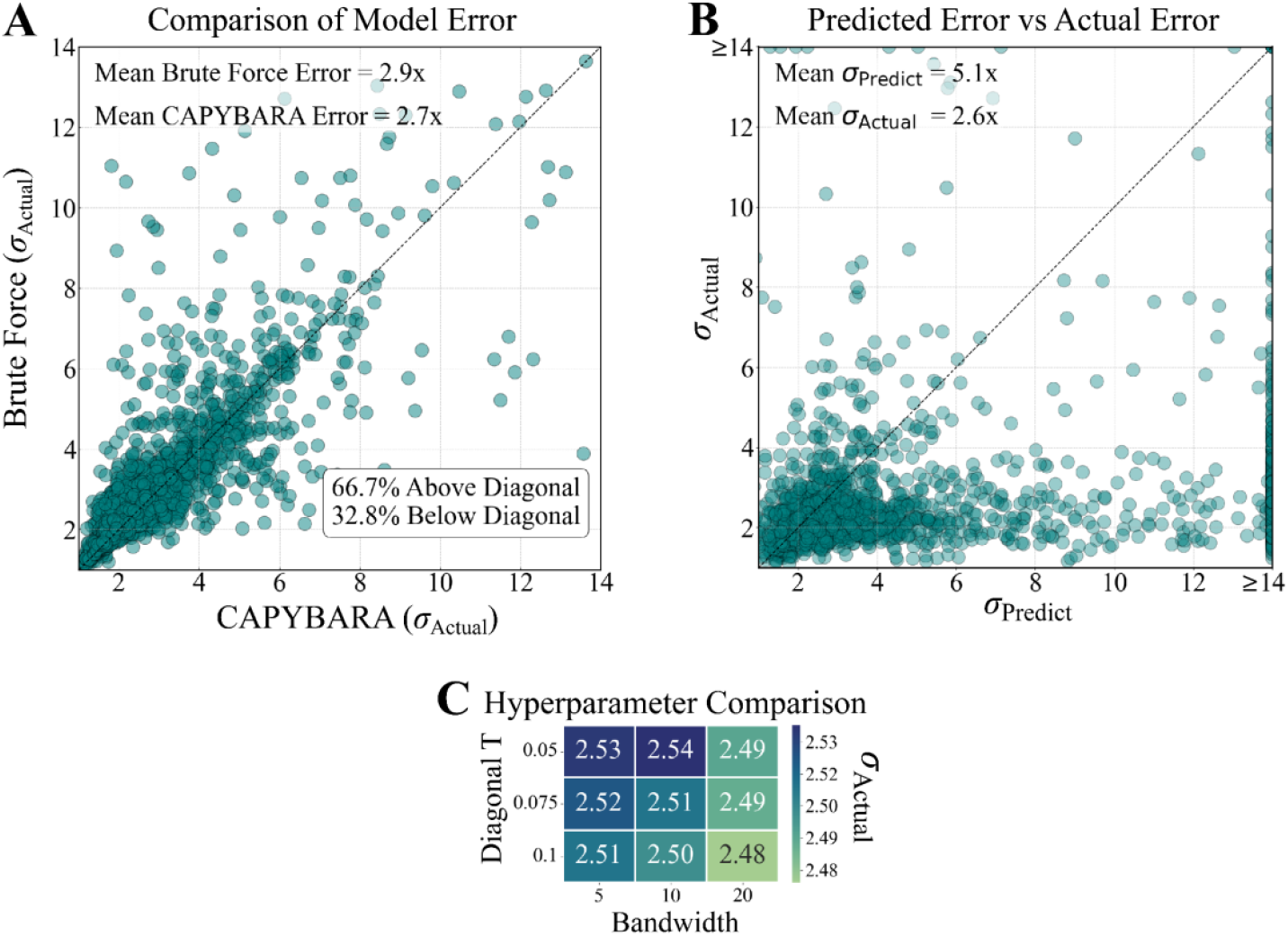
CAPYBARA achieves lower prediction error than brute force approaches and allows for predictive uncertainty estimation. (A) Comparison of fold-error for pairwise models generated by brute-force selection (running ridge regression on five randomly selected viruses, repeating 50 times to find the best five viruses) versus CAPYBARA (runs RFM a single time to identify the most predictive features and then ridge regression). Each point represents an overlapping virus between each dataset pair. More points lie above the diagonal and the average error is slightly smaller along the *x*-axis, with both traits indicating better performance with CAPYBARA. (B) Predicted versus actual error across all datasets using CAPYBARA, with each point representing all measurements for one virus in one study. We expect the predicted error to represent an upper bound, worst case error (*σ*_Actual_≲*σ*_Predict_), which is satisfied in the vast majority of cases. (C) Heatmap of mean *σ*_Actual_ across all dataset pairs for different hyperparameter settings for the diagonal threshold and bandwidth in RFM, showing nearly comparable prediction accuracy across all parameter choices.

## References

1. Andrews, S.F., Huang, Y., Kaur, K., Popova, L.I., Ho, I.Y., Pauli, N.T., Dunand, C.J.H., Taylor, W.M., Lim, S., Huang, M., et al. (2015). Immune history profoundly affects broadly protective B cell responses to influenza. Science Translational Medicine 7, 316ra192–316ra311. 10.1126/scitranslmed.aad0522.

2. Gostic, K.M., Ambrose, M., Worobey, M., and Lloyd-Smith, J.O. (2016). Potent protection against H5N1 and H7N9 influenza via childhood hemagglutinin imprinting. Science 354, 722–726. 10.1126/science.aag1322.

3. Hopping, A.M., McElhaney, J., Fonville, J.M., Powers, D.C., Beyer, W.E.P., and Smith, D.J. (2016). The confounded effects of age and exposure history in response to influenza vaccination. Vaccine 34, 540–546. 10.1016/j.vaccine.2015.11.058.

4. Vinh, D.N., Nhat, N.T.D., De Bruin, E., Vy, N.H.T., Thao, T.T.N., Phuong, H.T., Anh, P.H., Todd, S., Quan, T.M., Thanh, N.T.L., et al. (2021). Age-seroprevalence curves for the multi-strain structure of influenza A virus. Nature Communications 12, 1–9. 10.1038/s41467-021-26948-8.

5. Fox, A., Carolan, L., Leung, V., Phuong, H.V.M., Khvorov, A., Auladell, M., Tseng, Y.Y., Thai, P.Q., Barr, I., Subbarao, K., et al. (2022). Opposing Effects of Prior Infection versus Prior Vaccination on Vaccine Immunogenicity against Influenza A(H3N2) Viruses. Viruses 14, 1–15. 10.3390/v14030470.

6. Loes, A.N., Tarabi, R.A.L., Huddleston, J., Touyon, L., Wong, S.S., Cheng, S.M.S., Leung, N.H.L., Hannon, W.W., Bedford, T., Cobey, S., et al. (2024). High-throughput sequencing-based neutralization assay reveals how repeated vaccinations impact titers to recent human H1N1 influenza strains. Journal of Virology 98, 1–28. 10.1128/jvi.00689-24.

7. Lessler, J., Riley, S., Read, J.M., Wang, S., Zhu, H., Smith, G.J., Guan, Y., Jiang, C.Q., and Cummings, D.A. (2012). Evidence for antigenic seniority in influenza A (H3N2) antibody responses in southern China. PLoS Pathog 8, e1002802. 10.1371/journal.ppat.1002802.

8. Henry, C., Zheng, N.Y., Huang, M., Cabanov, A., Rojas, K.T., Kaur, K., Andrews, S.F., Palm, A.E., Chen, Y.Q., Li, Y., et al. (2019). Influenza Virus Vaccination Elicits Poorly Adapted B Cell Responses in Elderly Individuals. Cell Host Microbe 25, 357–366 e356. 10.1016/j.chom.2019.01.002.

9. Gouma, S., Kim, K., Weirick, M.E., Gumina, M.E., Branche, A., Topham, D.J., Martin, E.T., Monto, A.S., Cobey, S., and Hensley, S.E. (2020). Middle-aged individuals may be in a perpetual state of H3N2 influenza virus susceptibility. Nat Commun 11, 4566. 10.1038/s41467-020-18465-x.

10. Brouwer, A.F., Balmaseda, A., Gresh, L., Patel, M., Ojeda, S., Schiller, A.J., Lopez, R., Webby, R.J., Nelson, M.I., Kuan, G., and Gordon, A. (2022). Birth cohort relative to an influenza A virus’s antigenic cluster introduction drives patterns of children’s antibody titers. PLoS Pathog 18, e1010317. 10.1371/journal.ppat.1010317.

11. Kim, K., Marcos, Gouma, S., Madison, Scott, and Cobey, S. (2024). Measures of Population Immunity Can Predict the Dominant Clade of Influenza A (H3N2) in the 2017–2018 Season and Reveal Age-Associated Differences in Susceptibility and Antibody-Binding Specificity. Influenza and Other Respiratory Viruses 18, 1–13. 10.1111/irv.70033.

12. Xie, H., Wan, X.-F., Ye, Z., Plant, E.P., Zhao, Y., Xu, Y., Li, X., Finch, C., Zhao, N., Kawano, T., et al. (2015). H3N2 Mismatch of 2014–15 Northern Hemisphere Influenza Vaccines and Head-to-head Comparison between Human and Ferret Antisera derived Antigenic Maps. Scientific Reports 5, 15279. 10.1038/srep15279.

13. Morris, D.H., Gostic, K.M., Pompei, S., Bedford, T., Luksza, M., Neher, R.A., Grenfell, B.T., Lassig, M., and McCauley, J.W. (2018). Predictive Modeling of Influenza Shows the Promise of Applied Evolutionary Biology. Trends Microbiol 26, 102–118. 10.1016/j.tim.2017.09.004.

14. Zhao, X., Fang, V.J., Ohmit, S.E., Monto, A.S., Cook, A.R., and Cowling, B.J. (2016). Quantifying Protection Against Influenza Virus Infection Measured by Hemagglutination-inhibition Assays in Vaccine Trials. Epidemiology 27, 143–151. 10.1097/EDE.0000000000000402.

15. Cowling, B.J., Lim, W.W., Perera, R., Fang, V.J., Leung, G.M., Peiris, J.S.M., and Tchetgen Tchetgen, E.J. (2019). Influenza Hemagglutination-inhibition Antibody Titer as a Mediator of Vaccine-induced Protection for Influenza B. Clin Infect Dis 68, 1713–1717. 10.1093/cid/ciy759.

16. Krammer, F. (2019). The human antibody response to influenza A virus infection and vaccination. Nat Rev Immunol 19, 383–397. 10.1038/s41577-019-0143-6.

17. Kucharski, A.J., Lessler, J., Read, J.M., Zhu, H., Jiang, C.Q., Guan, Y., Cummings, D.A.T., and Riley, S. (2015). Estimating the Life Course of Influenza A(H3N2) Antibody Responses from Cross-Sectional Data. PLOS Biology 13, e1002082. 10.1371/journal.pbio.1002082.

18. Kucharski, A.J., Lessler, J., Cummings, D.A.T., and Riley, S. (2018). Timescales of influenza A/H3N2 antibody dynamics. PLoS Biol 16, e2004974. 10.1371/journal.pbio.2004974.

19. Stacey, H., Carlock, M.A., Allen, J.D., Hanley, H.B., Crotty, S., Ross, T.M., and Einav, T. (2025). Leveraging pre-vaccination antibody titres across multiple influenza H3N2 variants to forecast the post-vaccination response. eBioMedicine 116, 105744. 10.1016/j.ebiom.2025.105744.

20. Lapedes, A., and Farber, R. (2001). The geometry of shape space: application to influenza. J Theor Biol 212, 57–69. 10.1006/jtbi.2001.2347.

21. Smith, D.J., Lapedes, A.S., de Jong, J.C., Bestebroer, T.M., Rimmelzwaan, G.F., Osterhaus, A.D., and Fouchier, R.A. (2004). Mapping the antigenic and genetic evolution of influenza virus. Science 305, 371–376. 10.1126/science.1097211.

22. Anderson, C.S., McCall, P.R., Stern, H.A., Yang, H., and Topham, D.J. (2018). Antigenic cartography of H1N1 influenza viruses using sequence-based antigenic distance calculation. BMC Bioinformatics 19, 51. 10.1186/s12859-018-2042-4.

23. Einav, T., and Cleary, B. (2022). Extrapolating missing antibody-virus measurements across serological studies. Cell Syst 13, 561–573 e565. 10.1016/j.cels.2022.06.001.

24. Radhakrishnan, A., Beaglehole, D., Pandit, P., and Belkin, M. (2024). Mechanism for feature learning in neural networks and backpropagation-free machine learning models. Science 383, 1461–1467. 10.1126/science.adi5639.

25. Einav, T., and Ma, R. (2023). Using interpretable machine learning to extend heterogeneous antibody-virus datasets. Cell Rep Methods 3, 100540. 10.1016/j.crmeth.2023.100540.

26. Fonville, J.M., Wilks, S.H., James, S.L., Fox, A., Ventresca, M., Aban, M., Xue, L., Jones, T.C., Le, N.M.H., Pham, Q.T., et al. (2014). Antibody landscapes after influenza virus infection or vaccination. Science 346, 996–1000. 10.1126/science.1256427.

27. Ertesvag, N.U., Cox, R.J., Lartey, S.L., Mohn, K.G., Brokstad, K.A., and Trieu, M.C. (2022). Seasonal influenza vaccination expands hemagglutinin-specific antibody breadth to older and future A/H3N2 viruses. NPJ Vaccines 7, 67. 10.1038/s41541-022-00490-0.

28. Hinojosa, M., Shepard, S.S., Chung, J.R., King, J.P., McLean, H.Q., Flannery, B., Belongia, E.A., and Levine, M.Z. (2021). Impact of Immune Priming, Vaccination, and Infection on Influenza A(H3N2) Antibody Landscapes in Children. J Infect Dis 224, 469–480. 10.1093/infdis/jiaa665.

29. Carlock, M.A., Allen, J.D., Hanley, H.B., and Ross, T.M. (2024). Longitudinal assessment of human antibody binding to hemagglutinin elicited by split-inactivated influenza vaccination over six consecutive seasons. PLOS ONE 19, e0301157. 10.1371/journal.pone.0301157.

30. Hay, J.A., Zhu, H., Jiang, C.Q., Kwok, K.O., Shen, R., Kucharski, A., Yang, B., Read, J.M., Lessler, J., Cummings, D.A.T., and Riley, S. (2024). Reconstructed influenza A/H3N2 infection histories reveal variation in incidence and antibody dynamics over the life course. PLOS Biology 22, e3002864. 10.1371/journal.pbio.3002864.

31. Welsh, F.C., Eguia, R.T., Lee, J.M., Haddox, H.K., Galloway, J., Van Vinh Chau, N., Loes, A.N., Huddleston, J., Yu, T.C., Quynh Le, M., et al. (2024). Age-dependent heterogeneity in the antigenic effects of mutations to influenza hemagglutinin. Cell Host & Microbe 32, 1–15. 10.1016/j.chom.2024.06.015.

32. Lane, A., Quach, H.Q., Ovsyannikova, I.G., Kennedy, R.B., Ross, T.M., and Einav, T. (2025). Characterizing the Short- and Long-Term Temporal Dynamics of Antibody Responses to Influenza Vaccination. medRxiv preprint. 10.1101/2025.02.26.25322965.

